# Monitoring SNAP and Food Insecurity After the One Big Beautiful Bill Act: Early Evidence from a National Longitudinal Cohort

**DOI:** 10.64898/2026.11.19.26368514

**Authors:** McKaylee Robertson, Jenna Sanborn, Kate Penrose, Josefina Nuñez Sahr, Sophia Arabadjis, Emily Silva, Madhura Rane, Angela Parcesepe, Denis Nash

## Abstract

The One Big Beautiful Bill Act (OBBBA), signed in July 2025, restricted eligibility for the Supplemental Nutrition Assistance Program (SNAP) and expanded its work requirement, while federal tracking of food insecurity was scaled back. Using a national, community-based longitudinal cohort, we examined changes in SNAP receipt and food insecurity between August 2025 and March 2026 among 844 adults with household income below 200% of the federal poverty guideline, comparing participants in states that had and had not begun enforcing the expanded work requirement. SNAP receipt fell from 40.4% to 38.4%, and the share screening positive for food insecurity rose from 48.1% to 51.2%, with no detectable difference by state implementation timing. Adults who discontinued SNAP were as likely to be food insecure (71.1% in March 2026) and as worried about affording food, housing, and utilities as those who continued SNAP, and few had moved above 200% of the poverty guideline. These early findings suggest that need persisted after SNAP ended. As enforcement expands, policy makers should consider measures that keep benefits flowing to eligible people, such as extended certification periods and simplified recertification, and should support longitudinal monitoring of food insecurity.

**TEASER TEXT:** Federal changes to the Supplemental Nutrition Assistance Program (SNAP) in 2025 tightened work requirements just as the government stopped tracking food insecurity. Following the same 844 low-income adults from August 2025 to March 2026, we found that half screened positive for food insecurity and that the share rose over time. People who left SNAP were just as likely to be food insecure, and just as worried about paying for food, housing, and utilities, as those who stayed on, suggesting that need did not end when benefits did.

## INTRODUCTION

Food insecurity, the condition of having uncertain or limited access to adequate food, increased in recent years.^1–3^ According to the most recent national estimates, 13.7% of U.S. households experienced food insecurity in 2024, a prevalence significantly higher than levels observed between 2016 and 2021.^1–3^ Estimates are higher among working-age adults and in more recent non-federal surveys.^4^ Food insecurity is associated with poorer physical and mental health^5,6^ and higher healthcare utilization and cost^7^. Disabled adults, single mothers, and low-income households disproportionately experience food insecurity.^2,8^ The Supplemental Nutrition Assistance Program (SNAP), the largest federal nutrition assistance program, provides monthly benefits to low-income households, and has been found to alleviate food insecurity^5,9^ so policies that restrict eligibility and discourage participation may worsen food insecurity.^10,11^

Signed into law in July 2025, the One Big Beautiful Bill Act (OBBBA) introduced substantial changes to SNAP, including expanded work-reporting requirements, reduced federal funding for state administration, and narrowed eligibility.^12–14^ Prior to OBBBA, SNAP already imposed general work requirements for adults aged 18–54, alongside additional work requirements and time limits for able-bodied adults without dependents (ABAWDs). Under the ABAWD rules, individuals who do not meet the applicable work requirements or qualify for an exemption can receive benefits for a maximum of 3 months within a 3-year period. OBBBA raised the maximum age subject to ABAWD time limit from 54 to 64 years, removed exemptions for groups including veterans and individuals experiencing homelessness, and restricted states’ ability to obtain waivers in areas with insufficient job opportunities.^12–14^ OBBBA also ended eligibility for many lawfully present immigrants, including refugees and asylees, and shifted administrative costs to states.^12–14^ Beyond terminations, these changes add complexity and administrative burden for households and states with less funding, which may delay or interrupt benefits for eligible people.

SNAP participation was declining before OBBBA because of the de-implementation of pandemic-era allotments and flexibilities and anticipatory churn (where the process of policy change itself may result in disenrollment).^15,16^ Most states (39) began monitoring ABAWD noncompliance in December 2025, and the earliest SNAP terminations were possible around March 2026.^17^ Nationally, SNAP participation fell by approximately 13% from enactment to June 2026, with declines in nearly every state.^18^ SNAP benefits were also disrupted during the federal government shutdown in late 2025 (Oct 1-Nov 12)^19^, and its impact on food security is unknown.

Prior research on SNAP work requirements has found that they reduced participation without increasing employment^20–23^ and increased reliance on emergency food assistance, such as food pantries.^24^ Most of this evidence comes from administrative caseloads and repeated cross-sectional surveys, which show how many people leave SNAP but not what happens to them afterward. Few studies have used individual-level longitudinal data spanning these policy changes. Federal data on these questions are also shrinking: the USDA ended its annual food security report after releasing 2024 data.^25^ Population-level panel surveys, such as the Survey of Income and Program Participation, capture past-month SNAP receipt, but data covering the 2026 enforcement period are not expected before mid-2028.^26^ Little is known about how food insecurity has changed or who has started or stopped receiving SNAP during early OBBBA implementation.

Using longitudinal data from a national and sociodemographically diverse cohort of US adults followed since 2020^27^, we examined SNAP receipt and food insecurity among low-income adults during early OBBBA implementation from August 2025 to March 2026. The analysis captures cumulative early implementation dynamics because OBBBA enactment coincided with a downward trend in SNAP participation, administrative and anticipatory churn, and a federal government shutdown. First, we examined changes from baseline (August 2025) to follow-up (March 2026) in SNAP receipt and food insecurity, along with employment (the stated goal of ABAWD work requirements) and other measures of financial precarity. Second, we compared changes between participants in states where the expanded ABAWD/work requirement SNAP terminations were possible by March 2026 with participants in states where they could not yet occur. Third, we described food insecurity and compared affordability worries of adults who discontinued or initiated SNAP with adults whose SNAP status did not change.

## METHODS

### Data Source

The CHASING COVID Cohort is a community-based sample of adults aged 18 years and older and residing in the United States or US territories who were enrolled in a prospective internet-based study between March 28, 2020 and August 21, 2020. Details of cohort recruitment and follow-up have been described elsewhere.^27^ Follow-up is ongoing. Participants recently completed assessments in August 2025, after OBBBA was signed but before most eligibility changes were applied, and in March 2026, when terminations under the expanded ABAWD time limit had just become possible in early-implementing states but could not yet occur in other states.^17,28^

The primary analytic sample included 844 participants with household income below 200% of the federal poverty guideline (FPG) in August 2025 who completed both assessments (Supplemental Figure 1). SNAP eligibility depends on gross income (generally ≤130% FPG)^29^, net income (≤100% FPG), and assets, but through broad-based categorical eligibility most states raise the gross income limit, often to 200% FPG.^30^ We used income below 200% FPG as a proxy for likely eligibility.^31^ Because eligibility rules vary by state and income can change during the year, we repeated analyses among all participants who completed both assessments (N = 3,406).

### SNAP Eligibility Based on Federal Poverty Guideline

As others have done,^31^ we considered participants to be likely eligible for SNAP if they had a household income below 200% FPG in August 2025.^32^ Each participant was assigned the midpoint of their household income category, which was divided by the 2025 HHS poverty guideline for their household size. Using the midpoint produces a lower poverty rate^33^ and should conservatively identify those <200% FPG.

### Research Ethics

The study protocol was approved by the Institutional Review Board of the City University of New York (CUNY) (Protocol 2020-0256). Participant consent was obtained at baseline and at periodic follow-up assessments.

### Primary Outcomes: SNAP Receipt and Food Insecurity

At the August 2025 and March 2026 assessments, participants were asked “During the past 12 months, have you received food stamps, also called SNAP, the Supplemental Nutrition Assistance Program on an EBT card?” Responses were dichotomized as Yes or No/Do not know at each assessment.

At the March 2026 assessment, participants reporting SNAP in the past 12 months were also asked “During the past 30 days, have you received food stamps, also called SNAP, the Supplemental Nutrition Assistance Program on an EBT card?” Responses were dichotomized as Yes or No/Do not know. Because our interest was in SNAP status at the March 2026 assessment relative to August 2025, we classified participants into four mutually exclusive groups based on their reported receipt at each assessment: *continued on SNAP* (receipt in the past 30 days in March 2026 and in the past 12 months in August 2025), *initiated SNAP* (receipt in the past 30 days in March 2026 and none in the past 12 months in August 2025), *discontinued SNAP* (receipt in the past 12 months in March 2026 but not in the past 30 days), and *not on SNAP* (no receipt in the past 12 months in March 2026). Participants who reported receipt in the 12 months before August 2025 but no receipt in the 12 months before March 2026 (n = 52) had stopped before March 2025 and were classified as not on SNAP. Item nonresponse was low (<1.5%) and classified as no receipt.

Food insecurity was measured at each assessment using the Hunger Vital Sign, a validated two-item screener.^34,35^ Adults were considered food insecure if they endorsed either of the following statements as often or sometimes true in the past 12 months: (1) “We worried whether our food would run out before we got money to buy more.” or (2) “The food we bought just didn’t last and we didn’t have money to get more.” Against the 18-item USDA Household Food Security Survey Module, this screener has 97% sensitivity and 83% specificity, so it classifies more households as food insecure than the full module.^34^

### Secondary Outcomes: Employment, Utility Hardship, Housing Instability, and Affordability Worries

Employment, utility hardship, and housing instability were assessed in August 2025 and March 2026. We categorized employment status at both assessments as currently employed (“Employed for wages” or “Self-employed”) or not (“Out of work for 1 year or more”, “Out of work for less than 1 year”, “A homemaker”, “A student”, “Retired”, or “Unable to work”).

We considered participants to have a utility hardship at each assessment if, in the past 12 months, they reported they could not pay their mortgage, rent, or utility bills or a utility company threatened to shut off services. We used a 10-item housing instability index at both assessments.^36^ We considered participants to have any housing instability if they endorsed any adverse indicator (across 8 yes/no items), reported moving two or more times in the past 6 months, or stated they were unlikely or very unlikely to be able to pay for housing.

At the March 2026 assessment only, we also asked participants whether they were currently worried about being able to afford rent or mortgage, food, or utilities. Participants who indicated they were somewhat or very worried were categorized as worried for each item. For all secondary binary outcomes, missing responses (<1.4% across assessments) were coded as No.

### Timing of State ABAWD Implementation

Supplemental Table 1 lists each state and its classification as early- or not-yet-implementing for the ABAWD work requirement under OBBBA as of March 2026.^17^ We used the ZIP code reported at the most recent (March 2026) assessment to classify participants, as that location would govern SNAP receipt. In the not-yet-implementing states, no participants could have lost SNAP benefits under the time limit by March 2026. We excluded participants living in Puerto Rico or other US territories (n = 6), where SNAP does not operate. Participants with missing state (n = 42; 12 with income below 200% FPG) were included in overall estimates but not in comparisons between state groups.

### Other Characteristics

We examined sociodemographics (age, gender, race/ethnicity, education, employment, marital status), having any child <18 in the household, being a single parent of a child <18, being the parent of a child <14, and having any disability (ACS 6-item disability scale).^37^ We classified adults as likely subject to the work requirement if, in August 2025, they were aged 18–64, had no child under 14 in the household, reported no disability, and were not pregnant; all others were classified as nominally exempt.

### Statistical Analysis

Analyses were run among the likely SNAP-eligible population. We repeated analyses among all participants who completed both assessments, regardless of income, since <200% FPG may miss some SNAP-eligible participants. As a sensitivity analysis, we constructed stabilized inverse probability of censoring weights (IPCW) for loss to follow-up at the March 2026 assessment. See Supplemental Appendix 1 for model details related to the IPCW.

We used generalized estimating equations (GEE) with a modified Poisson model to generate relative risks (RR) comparing each outcome at follow-up (March 2026) with baseline (August 2025) overall and separately in early- and not-yet-implementing states. As noted above, these estimates describe change during the first months of OBBBA implementation and cannot be attributed to any single provision, particularly because SNAP participation was already declining before OBBBA. Of these concurrent changes, the timing of ABAWD enforcement is the one we could classify by state. We therefore compared change between state groups using a model with assessment wave, state group, and their interaction. The exponentiated interaction term is the ratio of RRs (early vs. not-yet-implementing).

For the four SNAP status groups, we described characteristics, food insecurity, and affordability worries. We compared affordability worries among groups that could make the same SNAP transition (e.g., discontinuing SNAP among those on SNAP and initiating among those not on SNAP). Specifically, we compared those who discontinued with those who continued SNAP, and those who initiated with those not on SNAP, using modified Poisson regression with robust standard errors. We also compared discontinuing and initiating SNAP between state groups. Because few participants discontinued or initiated, these analyses were descriptive and unadjusted.

Analyses were conducted using SAS 9.4 and R (version 4.5.1).

## RESULTS

A total of 3,770 participants completed the August 2025 assessment, and 3,412 (91%) participants completed the March 2026 follow-up. After excluding 6 participants living in US territories, the main analytic sample included 844 participants with a household income below 200% of the FPG (Supplemental Figure 1). Of these, 524 (62%) resided in early-implementing states and 308 (36%) resided in not-yet-implementing states, which had not begun enforcing the ABAWD time limit as of the March 2026 assessment; 12 (1%) had missing state (Table 1). Compared with participants in not-yet-implementing states, those in early-implementing states were more likely to be female (71.8% vs. 56.5%), Black non-Hispanic (20.8% vs. 13.0%), employed (51.5% vs. 44.2%), and to have a child under 18 in the household (36.5% vs. 26.6%), and less likely to be college graduates (26.9% vs. 41.6%). Age, disability (36.8% vs. 35.7%), and the share likely subject to the work requirement (37.6% vs. 40.3%) did not differ (Table 1).

**Table 1.**
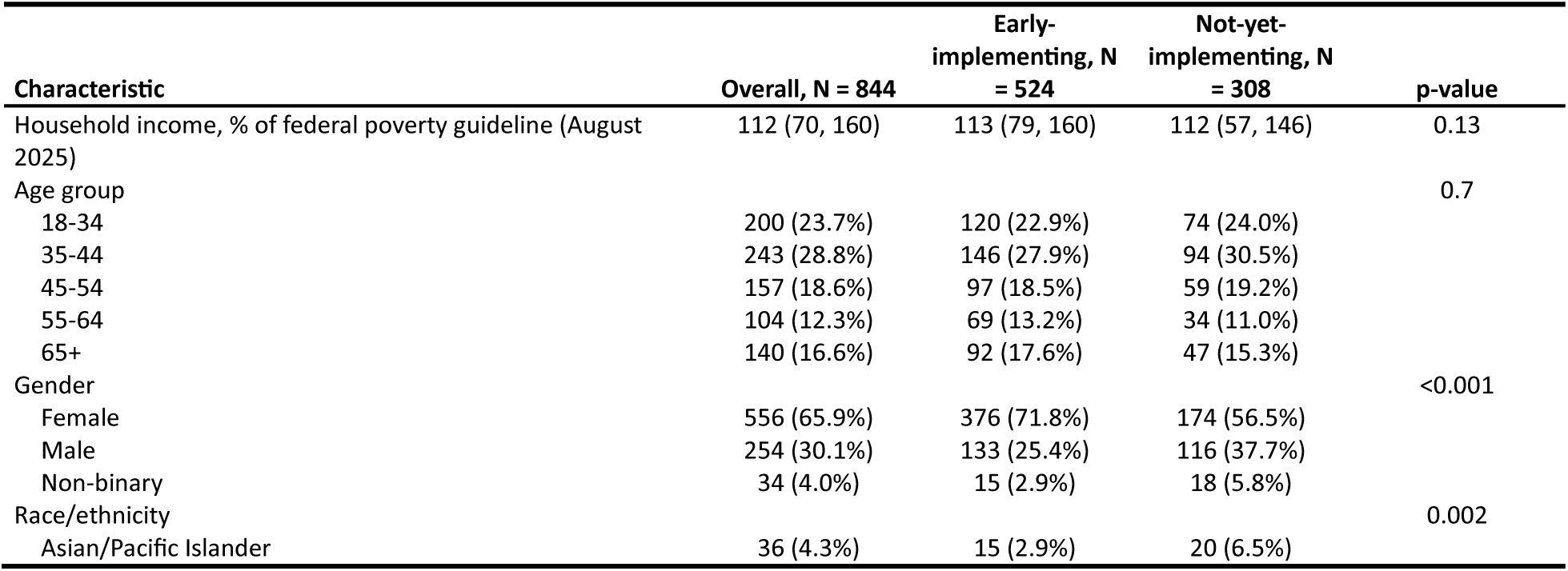
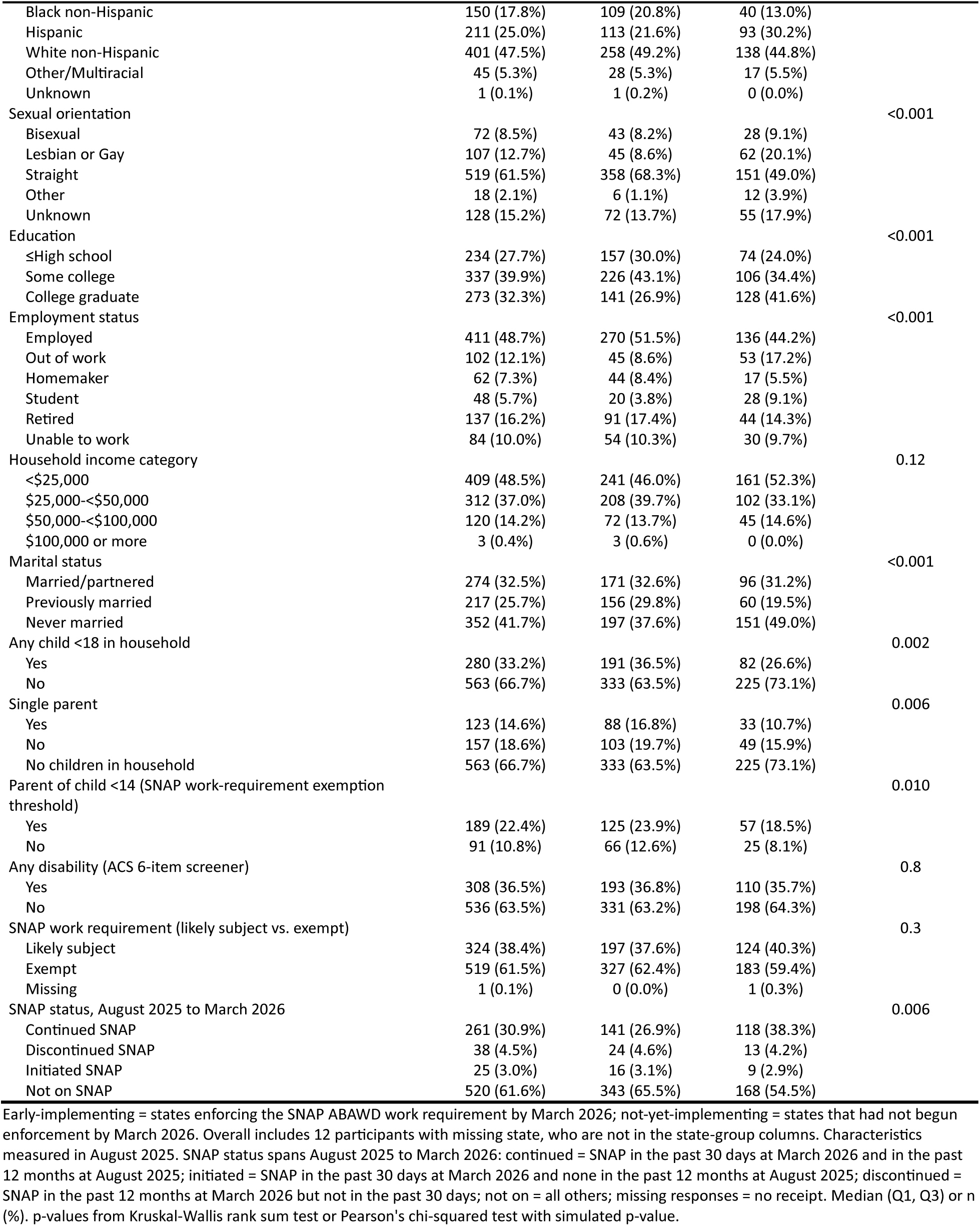
Characteristics of Adults with Household Income <200% FPG, by State ABAWD Implementation Timing.

| **Characteristic** | **Overall, N = 844** | **Early-implementing, N = 524** | **Not-yet-implementing, N = 308** | **p-value** |
|---|---|---|---|---|
| Household income, % of federal poverty guideline (August 2025) | 112 (70, 160) | 113 (79, 160) | 112 (57, 146) | 0.13 |
| Age group |  |  |  | 0.7 |
| 18-34 | 200 (23.7%) | 120 (22.9%) | 74 (24.0%) |  |
| 35-44 | 243 (28.8%) | 146 (27.9%) | 94 (30.5%) |  |
| 45-54 | 157 (18.6%) | 97 (18.5%) | 59 (19.2%) |  |
| 55-64 | 104 (12.3%) | 69 (13.2%) | 34 (11.0%) |  |
| 65+ | 140 (16.6%) | 92 (17.6%) | 47 (15.3%) |  |
| Gender |  |  |  | <0.001 |
| Female | 556 (65.9%) | 376 (71.8%) | 174 (56.5%) |  |
| Male | 254 (30.1%) | 133 (25.4%) | 116 (37.7%) |  |
| Non-binary | 34 (4.0%) | 15 (2.9%) | 18 (5.8%) |  |
| Race/ethnicity |  |  |  | 0.002 |
| Asian/Pacific Islander | 36 (4.3%) | 15 (2.9%) | 20 (6.5%) |  |
| Black non-Hispanic | 150 (17.8%) | 109 (20.8%) | 40 (13.0%) |  |
| Hispanic | 211 (25.0%) | 113 (21.6%) | 93 (30.2%) |  |
| White non-Hispanic | 401 (47.5%) | 258 (49.2%) | 138 (44.8%) |  |
| Other/Multiracial | 45 (5.3%) | 28 (5.3%) | 17 (5.5%) |  |
| Unknown | 1 (0.1%) | 1 (0.2%) | 0 (0.0%) |  |
| Sexual orientation |  |  |  | <0.001 |
| Bisexual | 72 (8.5%) | 43 (8.2%) | 28 (9.1%) |  |
| Lesbian or Gay | 107 (12.7%) | 45 (8.6%) | 62 (20.1%) |  |
| Straight | 519 (61.5%) | 358 (68.3%) | 151 (49.0%) |  |
| Other | 18 (2.1%) | 6 (1.1%) | 12 (3.9%) |  |
| Unknown | 128 (15.2%) | 72 (13.7%) | 55 (17.9%) |  |
| Education |  |  |  | <0.001 |
| ≤High school | 234 (27.7%) | 157 (30.0%) | 74 (24.0%) |  |
| Some college | 337 (39.9%) | 226 (43.1%) | 106 (34.4%) |  |
| College graduate | 273 (32.3%) | 141 (26.9%) | 128 (41.6%) |  |
| Employment status |  |  |  | <0.001 |
| Employed | 411 (48.7%) | 270 (51.5%) | 136 (44.2%) |  |
| Out of work | 102 (12.1%) | 45 (8.6%) | 53 (17.2%) |  |
| Homemaker | 62 (7.3%) | 44 (8.4%) | 17 (5.5%) |  |
| Student | 48 (5.7%) | 20 (3.8%) | 28 (9.1%) |  |
| Retired | 137 (16.2%) | 91 (17.4%) | 44 (14.3%) |  |
| Unable to work | 84 (10.0%) | 54 (10.3%) | 30 (9.7%) |  |
| Household income category |  |  |  | 0.12 |
| <\$25,000 | 409 (48.5%) | 241 (46.0%) | 161 (52.3%) |  |
| \$25,000-<\$50,000 | 312 (37.0%) | 208 (39.7%) | 102 (33.1%) |  |
| \$50,000-<\$100,000 | 120 (14.2%) | 72 (13.7%) | 45 (14.6%) |  |
| \$100,000 or more | 3 (0.4%) | 3 (0.6%) | 0 (0.0%) |  |
| Marital status |  |  |  | <0.001 |
| Married/partnered | 274 (32.5%) | 171 (32.6%) | 96 (31.2%) |  |
| Previously married | 217 (25.7%) | 156 (29.8%) | 60 (19.5%) |  |
| Never married | 352 (41.7%) | 197 (37.6%) | 151 (49.0%) |  |
| Any child <18 in household |  |  |  | 0.002 |
| Yes | 280 (33.2%) | 191 (36.5%) | 82 (26.6%) |  |
| No | 563 (66.7%) | 333 (63.5%) | 225 (73.1%) |  |
| Single parent |  |  |  | 0.006 |
| Yes | 123 (14.6%) | 88 (16.8%) | 33 (10.7%) |  |
| No | 157 (18.6%) | 103 (19.7%) | 49 (15.9%) |  |
| No children in household | 563 (66.7%) | 333 (63.5%) | 225 (73.1%) |  |
| Parent of child <14 (SNAP work-requirement exemption threshold) |  |  |  | 0.010 |
| Yes | 189 (22.4%) | 125 (23.9%) | 57 (18.5%) |  |
| No | 91 (10.8%) | 66 (12.6%) | 25 (8.1%) |  |
| Any disability (ACS 6-item screener) |  |  |  | 0.8 |
| Yes | 308 (36.5%) | 193 (36.8%) | 110 (35.7%) |  |
| No | 536 (63.5%) | 331 (63.2%) | 198 (64.3%) |  |
| SNAP work requirement (likely subject vs. exempt) |  |  |  | 0.3 |
| Likely subject | 324 (38.4%) | 197 (37.6%) | 124 (40.3%) |  |
| Exempt | 519 (61.5%) | 327 (62.4%) | 183 (59.4%) |  |
| Missing | 1 (0.1%) | 0 (0.0%) | 1 (0.3%) |  |
| SNAP status, August 2025 to March 2026 |  |  |  | 0.006 |
| Continued SNAP | 261 (30.9%) | 141 (26.9%) | 118 (38.3%) |  |
| Discontinued SNAP | 38 (4.5%) | 24 (4.6%) | 13 (4.2%) |  |
| Initiated SNAP | 25 (3.0%) | 16 (3.1%) | 9 (2.9%) |  |
| Not on SNAP | 520 (61.6%) | 343 (65.5%) | 168 (54.5%) |  |

Early-implementing = states enforcing the SNAP ABAWD work requirement by March 2026; not-yet-implementing = states that had not begun enforcement by March 2026. Overall includes 12 participants with missing state, who are not in the state-group columns. Characteristics measured in August 2025. SNAP status spans August 2025 to March 2026: continued = SNAP in the past 30 days at March 2026 and in the past 12 months at August 2025; initiated = SNAP in the past 30 days at March 2026 and none in the past 12 months at August 2025; discontinued = SNAP in the past 12 months at March 2026 but not in the past 30 days; not on = all others; missing responses = no receipt. Median (Q1, Q3) or n (%). p-values from Kruskal-Wallis rank sum test or Pearson's chi-squared test with simulated p-value.

### Change in Past 12-Month Outcomes, August 2025 to March 2026

Overall, SNAP receipt in the past 12 months decreased from 40.4% to 38.4% between August 2025 and March 2026 (RR 0.95, 95% CI 0.90–1.00), and past 12-month food insecurity increased from 48.1% to 51.2% (RR 1.06, 95% CI 1.01–1.12) among adults under 200% FPG (Table 2). SNAP receipt declined slightly in both early-implementing (36.1% to 34.5%; RR 0.96, 95% CI 0.88–1.04) and not-yet-implementing states (48.4% to 45.5%; RR 0.94, 95% CI 0.87–1.01), with no detectable difference between state groups (ratio of RRs 1.02, 95% CI 0.91–1.14). Food insecurity rose in both groups (early-implementing: 50.6% to 53.1% and not-yet-implementing: 44.5% to 48.4%), with no detectable difference between them (ratio of RRs 0.96, 95% CI 0.85–1.09).

**Table 2.** Change in Past 12-Month Outcomes from August 2025 to March 2026 Overall and by State ABAWD Implementation Timing, Among Adults with Household Income <200% FPG (N = 844)

| Outcome | State group | N | Aug 2025, n (%) | Mar 2026, n (%) | RR (95% CI) | RR, IPCW-weighted (95% CI) | Ratio of RRs, early- vs. not-yet-implementing (95% CI) | Ratio of RRs, IPCW-weighted (95% CI) |
|---|---|---|---|---|---|---|---|---|
| **Household income <200% FPG** |  |  |  |  |  |  |  |  |
| SNAP receipt | Early-implementing | 524 | 189 (36.1%) | 181 (34.5%) | 0.96 (0.88–1.04) | 0.96 (0.88–1.04) | 1.02 (0.91–1.14) | 1.02 (0.91–1.14) |
|  | Not-yet-implementing | 308 | 149 (48.4%) | 140 (45.5%) | 0.94 (0.87–1.01) | 0.94 (0.87–1.01) |  |  |
|  | **Overall** | 844 | 341 (40.4%) | 324 (38.4%) | **0.95 (0.90–1.00)** | **0.95 (0.90–1.00)** |  |  |
| Food insecurity | Early-implementing | 524 | 265 (50.6%) | 278 (53.1%) | 1.05 (0.98–1.12) | 1.04 (0.98–1.11) | 0.96 (0.85–1.09) | 0.96 (0.86–1.09) |
|  | Not-yet-implementing | 308 | 137 (44.5%) | 149 (48.4%) | 1.09 (0.98–1.21) | 1.08 (0.98–1.20) |  |  |
|  | **Overall** | 844 | 406 (48.1%) | 432 (51.2%) | **1.06 (1.01–1.12)** | **1.06 (1.00–1.12)** |  |  |
| Employed | Early-implementing | 524 | 270 (51.5%) | 275 (52.5%) | 1.02 (0.97–1.07) | 1.02 (0.97–1.07) | 0.98 (0.88–1.08) | 0.98 (0.88–1.08) |
|  | Not-yet-implementing | 308 | 136 (44.2%) | 142 (46.1%) | 1.04 (0.96–1.14) | 1.04 (0.96–1.14) |  |  |
|  | **Overall** | 844 | 411 (48.7%) | 423 (50.1%) | 1.03 (0.98–1.08) | 1.03 (0.98–1.08) |  |  |
| Any utility/bill hardship | Early-implementing | 524 | 181 (34.5%) | 156 (29.8%) | **0.86 (0.77–0.97)** | **0.86 (0.77–0.97)** | 0.87 (0.72–1.06) | 0.87 (0.71–1.06) |
|  | Not-yet-implementing | 308 | 94 (30.5%) | 93 (30.2%) | 0.99 (0.84–1.16) | 0.99 (0.85–1.16) |  |  |
|  | **Overall** | 844 | 276 (32.7%) | 251 (29.7%) | **0.91 (0.83–1.00)** | **0.91 (0.83–1.00)** |  |  |
| Housing instability | Early-implementing | 524 | 279 (53.2%) | 282 (53.8%) | 1.01 (0.93–1.09) | 1.01 (0.93–1.09) | 1.06 (0.93–1.20) | 1.06 (0.93–1.20) |
|  | Not-yet-implementing | 308 | 178 (57.8%) | 170 (55.2%) | 0.96 (0.87–1.05) | 0.95 (0.87–1.05) |  |  |
|  | **Overall** | 844 | 464 (55.0%) | 459 (54.4%) | 0.99 (0.93–1.05) | 0.99 (0.93–1.05) |  |  |

Income group defined in August 2025. Overall includes 12 participants with missing state, who are not in the state-group rows. Modified Poisson generalized estimating equations (log link, exchangeable correlation, clustered on participant). RR = March 2026 vs. August 2025 within each state group. Ratio of RRs = wave × state-group interaction (difference-in-differences on the ratio scale), reference = not-yet-implementing states. IPCW = stabilized inverse probability of censoring weights for loss to follow-up between assessments (truncated at the 1st and 99th percentiles; Supplemental Appendix 1); IPCW-weighted RRs and ratios of RRs are from the same models fit with these weights. SNAP receipt = past 12 months; food insecurity = Hunger Vital Sign.

Employment and housing instability did not change. Utility or bill hardship over the past 12 months declined in early-implementing states (34.5% to 29.8%; RR 0.86, 95% CI 0.77–0.97) but not in not-yet-implementing states, but the difference between state groups was not statistically significant (ratio of RRs 0.87, 95% CI 0.72–1.06).

### Continuing, Discontinuing, and Initiating SNAP, Past 30 Days March 2026

Among the 844 participants with income below 200% FPG, 324 (38.4%) were on SNAP in the 12 months before March 2026: 261 (30.9%) continued SNAP between August 2025 and March 2026, 38 (4.5%) discontinued SNAP, and 25 (3.0%) initiated SNAP (Figure 1). Among participants with SNAP in the 12 months before March 2026, discontinuing SNAP was more common in early-implementing states (24/181 [13.3%]) than in not-yet-implementing states (13/140 [9.3%]). Among those with no SNAP in the 12 months before August 2025, initiating SNAP was less common in early-versus not-yet-implementing states (16/335 [4.8%] versus 9/159 [5.7%], respectively). However, estimates were imprecise due to small sample size (discontinuing [RR 1.43, 95% CI 0.75–2.70] and initiating SNAP [RR 0.84, 95% CI 0.38–1.87]) (Supplemental Table 2).

**Figure 1.**
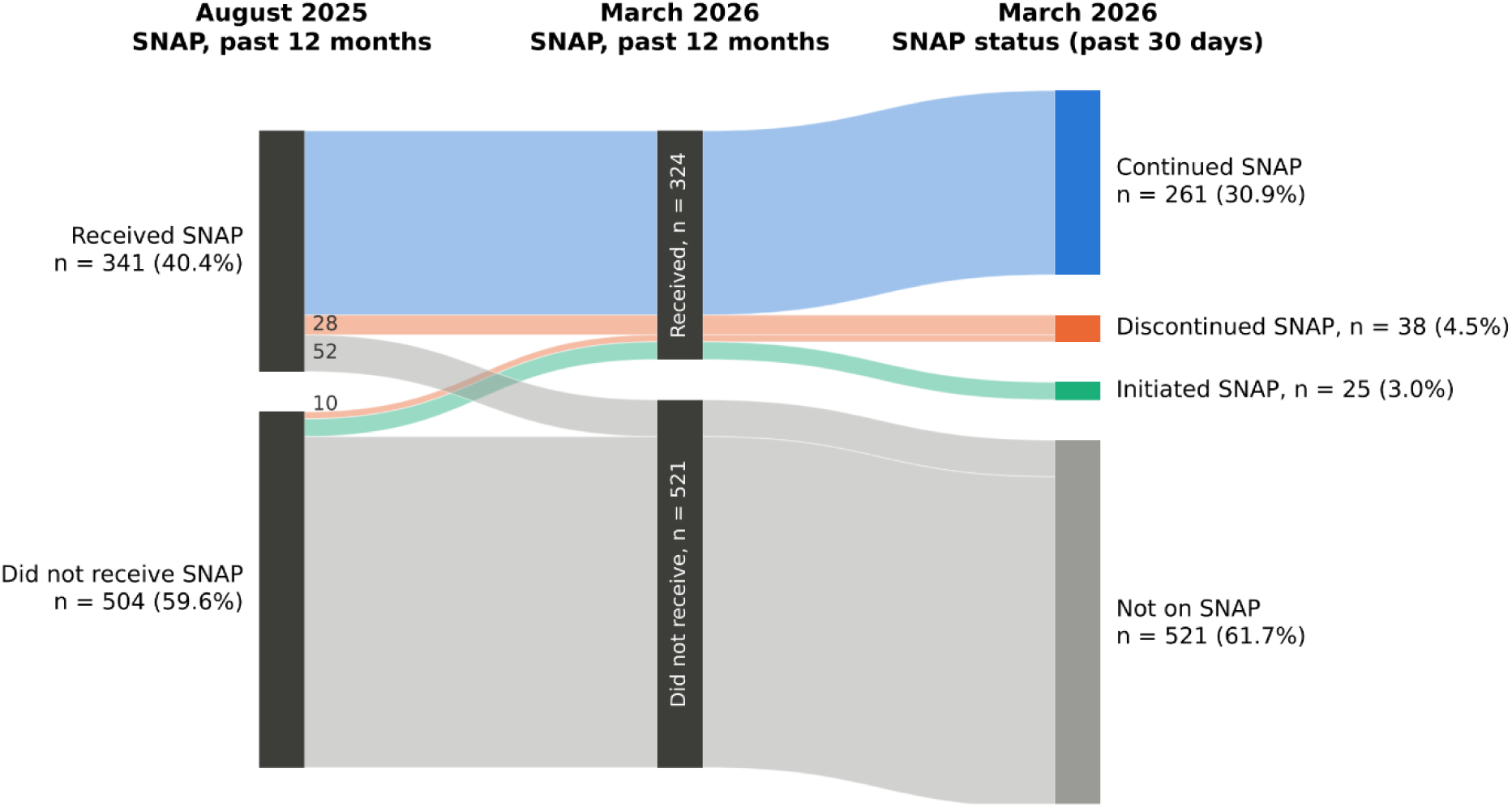
SNAP transitions from August 2025 to March 2026.

Compared with participants who continued SNAP, those who discontinued were more often employed in August 2025 (65.8% vs. 34.5%) and married/partnered (50.0% vs. 19.9%), and less often had household income below $25,000 (44.7% vs. 73.6%) or were aged 65 or older (0% vs. 17.2%) (Table 3A; Supplemental Table 3). Compared with participants not on SNAP, those who initiated SNAP were more often employed (68.0% vs. 53.7%) and less often college graduates (16.0% vs. 37.7%) or living with a disability (16.0% vs. 31.0%).

**Table 3.**
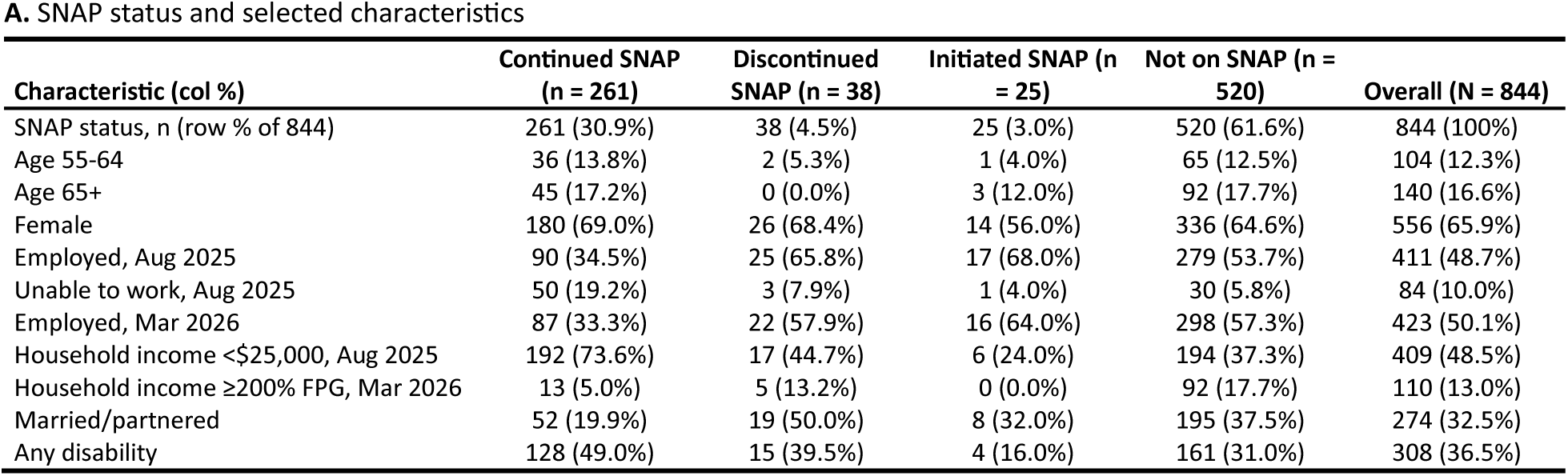
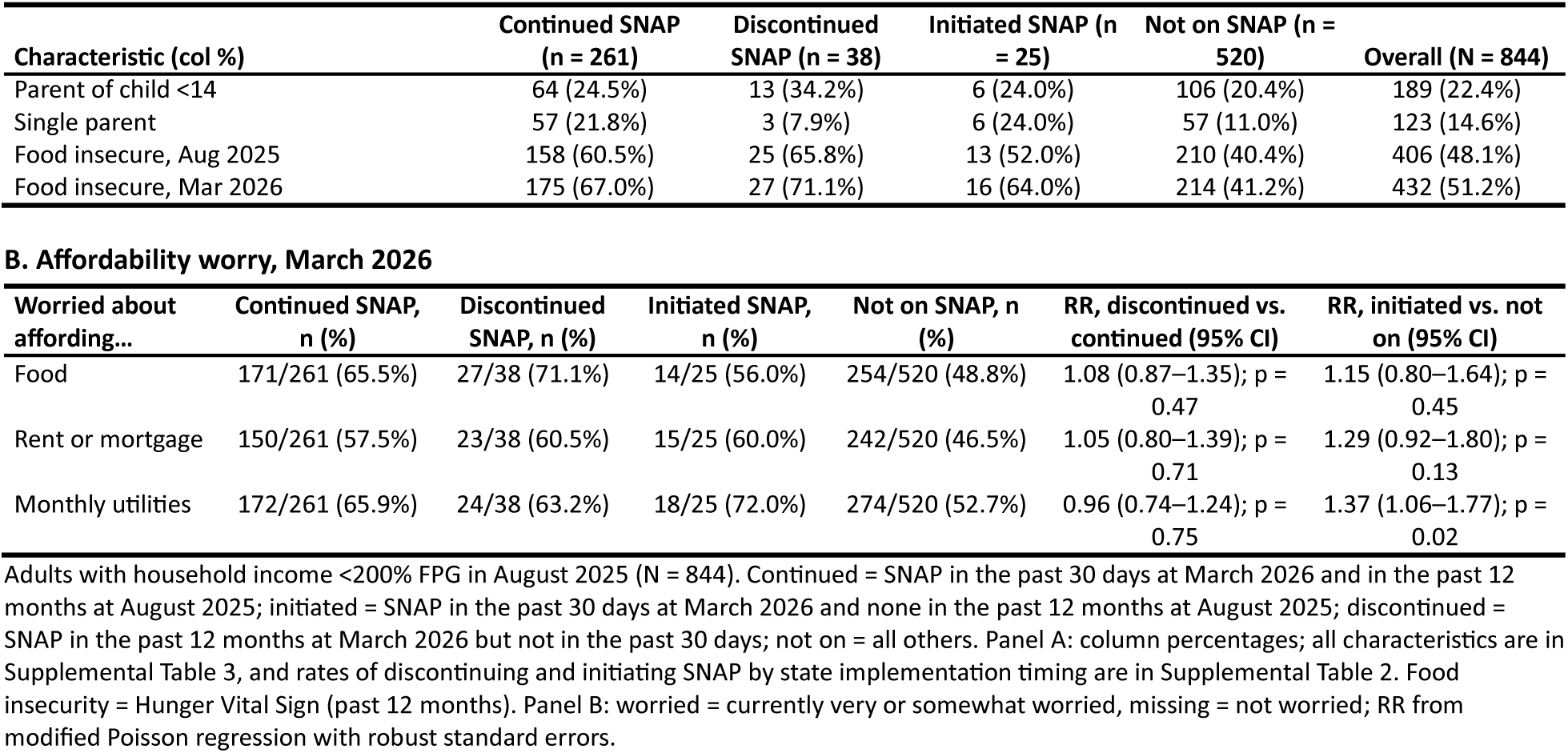
Who Continued, Discontinued or Initiated SNAP Between August 2025 and March 2026, and Food Insecurity and Affordability Worry by SNAP Status, Among Adults With Household Income <200% FPG

| **A.** SNAP status and selected characteristics |  |  |  |  |  |  |
|---|---|---|---|---|---|---|
| **Characteristic (col %)** | **Continued SNAP (n = 261)** | **Discontinued SNAP (n = 38)** | **Initiated SNAP (n = 25)** | **Not on SNAP (n = 520)** | **Overall (N = 844)** |  |
| SNAP status, n (row % of 844) | 261 (30.9%) | 38 (4.5%) | 25 (3.0%) | 520 (61.6%) | 844 (100%) |  |
| Age 55-64 | 36 (13.8%) | 2 (5.3%) | 1 (4.0%) | 65 (12.5%) | 104 (12.3%) |  |
| Age 65+ | 45 (17.2%) | 0 (0.0%) | 3 (12.0%) | 92 (17.7%) | 140 (16.6%) |  |
| Female | 180 (69.0%) | 26 (68.4%) | 14 (56.0%) | 336 (64.6%) | 556 (65.9%) |  |
| Employed, Aug 2025 | 90 (34.5%) | 25 (65.8%) | 17 (68.0%) | 279 (53.7%) | 411 (48.7%) |  |
| Unable to work, Aug 2025 | 50 (19.2%) | 3 (7.9%) | 1 (4.0%) | 30 (5.8%) | 84 (10.0%) |  |
| Employed, Mar 2026 | 87 (33.3%) | 22 (57.9%) | 16 (64.0%) | 298 (57.3%) | 423 (50.1%) |  |
| Household income <\$25,000, Aug 2025 | 192 (73.6%) | 17 (44.7%) | 6 (24.0%) | 194 (37.3%) | 409 (48.5%) |  |
| Household income ≥200% FPG, Mar 2026 | 13 (5.0%) | 5 (13.2%) | 0 (0.0%) | 92 (17.7%) | 110 (13.0%) |  |
| Married/partnered | 52 (19.9%) | 19 (50.0%) | 8 (32.0%) | 195 (37.5%) | 274 (32.5%) |  |
| Any disability | 128 (49.0%) | 15 (39.5%) | 4 (16.0%) | 161 (31.0%) | 308 (36.5%) |  |
| **Characteristic (col %)** | **Continued SNAP (n = 261)** | **Discontinued SNAP (n = 38)** | **Initiated SNAP (n = 25)** | **Not on SNAP (n = 520)** | **Overall (N = 844)** |  |
| Parent of child <14 | 64 (24.5%) | 13 (34.2%) | 6 (24.0%) | 106 (20.4%) | 189 (22.4%) |  |
| Single parent | 57 (21.8%) | 3 (7.9%) | 6 (24.0%) | 57 (11.0%) | 123 (14.6%) |  |
| Food insecure, Aug 2025 | 158 (60.5%) | 25 (65.8%) | 13 (52.0%) | 210 (40.4%) | 406 (48.1%) |  |
| Food insecure, Mar 2026 | 175 (67.0%) | 27 (71.1%) | 16 (64.0%) | 214 (41.2%) | 432 (51.2%) |  |
| **B. Affordability worry, March 2026** |  |  |  |  |  |  |
| **Worried about affording…** | **Continued SNAP, n (%)** | **Discontinued SNAP, n (%)** | **Initiated SNAP, n (%)** | **Not on SNAP, n (%)** | **RR, discontinued vs. continued (95% CI)** | **RR, initiated vs. not on (95% CI)** |
| Food | 171/261 (65.5%) | 27/38 (71.1%) | 14/25 (56.0%) | 254/520 (48.8%) | 1.08 (0.87–1.35); p = 0.47 | 1.15 (0.80–1.64); p = 0.45 |
| Rent or mortgage | 150/261 (57.5%) | 23/38 (60.5%) | 15/25 (60.0%) | 242/520 (46.5%) | 1.05 (0.80–1.39); p = 0.71 | 1.29 (0.92–1.80); p = 0.13 |
| Monthly utilities | 172/261 (65.9%) | 24/38 (63.2%) | 18/25 (72.0%) | 274/520 (52.7%) | 0.96 (0.74–1.24); p = 0.75 | 1.37 (1.06–1.77); p = 0.02 |

Adults with household income <200% FPG in August 2025 (N = 844). Continued = SNAP in the past 30 days at March 2026 and in the past 12 months at August 2025; initiated = SNAP in the past 30 days at March 2026 and none in the past 12 months at August 2025; discontinued = SNAP in the past 12 months at March 2026 but not in the past 30 days; not on = all others. Panel A: column percentages; all characteristics are in Supplemental Table 3, and rates of discontinuing and initiating SNAP by state implementation timing are in Supplemental Table 2. Food insecurity = Hunger Vital Sign (past 12 months). Panel B: worried = currently very or somewhat worried, missing = not worried; RR from modified Poisson regression with robust standard errors.

Food insecurity over the past 12 months was higher in March 2026 for those who continued, discontinued or initiated SNAP (64.0%–71.1%) than for those not on SNAP (41.2%) (Table 3A). Affordability worries in March 2026 were also higher among those who continued, discontinued, or initiated SNAP than for those not on SNAP (worried about food: 56.0%–71.1% versus 48.8%; rent/mortgage: 57.5%–60.5% versus 46.5%; monthly utilities: 63.2%–72.0% versus 52.7%) (Table 3B). Compared with those not on SNAP, those who initiated SNAP were more often worried about affording utilities (72.0% vs. 52.7%; RR 1.37, 95% CI 1.06–1.77). Affordability worries did not differ significantly between those who discontinued and those who continued SNAP.

Among all 3,406 participants regardless of income, SNAP receipt declined (14.5% to 13.6%; RR 0.94, 95% CI 0.89–0.99) and food insecurity changed little (22.2% to 23.3%; RR 1.05, 95% CI 1.00–1.10), with no difference between state groups for any outcome (Supplemental Table 4). RRs for discontinuing and initiating by state ABAWD timing were in the same direction and also not significant among all 3,406 adults regardless of income (Supplemental Table 5A). In exploratory analyses, change in SNAP receipt and food insecurity did not differ between adults likely subject to the work requirement and exempt adults across state groups (Supplemental Table 6).

## DISCUSSION

In the first months of OBBBA implementation, approximately half of adults under 200% FPG screened positive for food insecurity, rising from 48% in August 2025 to 51% in March 2026, while SNAP receipt declined from 40% to 38%. While the magnitude of the decline was modest, such declines represent a large number of households at a national scale. With the USDA food security survey ended and NHIS no longer tracking food insecurity,^25,38^ longitudinal cohorts may be among the few sources to monitor food insecurity as changes to SNAP take effect. These data establish an early within-cohort baseline for monitoring food insecurity as individuals’ SNAP status changes and OBBBA implementation expands.

Food insecurity and SNAP receipt did not detectably differ between states that were and were not yet implementing SNAP’s new work requirement, which is expected at this early stage. In most early-implementing states, months began counting toward the time limit on December 1, 2025, so March 2026 was among the first months in which SNAP terminations were possible. The relative decline in SNAP receipt in our cohort (5%) was similar to the national decline in average monthly participation over the same 12-month windows based on administrative counts (approximately 5%: September 2024-August 2025 versus April 2025-March 2026).^39^ SNAP receipt also declined in not-yet-implementing states, consistent with the national decline in participation that began before OBBBA.^15,16^ Because the August 2025 (pre-enforcement) assessment followed enactment, participants in both state groups may already have been leaving or not renewing SNAP before any terminations were possible. New rules and reporting requirements may discourage eligible people from enrolling or recertifying, especially when benefits seem uncertain or not worth the effort. Such effects would move both state groups in the same direction and the estimate toward the null.

Food insecurity was higher among participants who interacted with SNAP (continued, discontinued or initiated) (64.0-71.1%) than those who were not on SNAP (41.2%). This likely reflects greater underlying need: recipients had greater need than non-participants at baseline (August 2025), even with SNAP assistance.^5,9^ Our food insecurity screener asks about the past 12 months, so March 2026 responses span the late-2025 interruption of SNAP benefits during the federal government shutdown, which affected only recipients. These findings extend cross-sectional evidence that SNAP participants had high needs before OBBBA.^31^

Adults who discontinued SNAP were as food insecure and as worried about affording food, rent, and utilities in March 2026 as those who continued on SNAP, and few (5 of 38) had incomes at or above 200% FPG by March 2026. Although they were more often employed, their need persisted after assistance ended. Some may have experienced income changes, but our data cannot establish why individuals stopped reporting SNAP receipt. Persistent need is expected under a policy in which eligibility depends on work hours and administrative compliance rather than on food need, and policies that restrict access or add administrative barriers may reduce participation without resolving the underlying need for food.

Adults who initiated SNAP were also food insecure (64.0% in March 2026) and more often worried about affording utilities than adults not on SNAP, consistent with SNAP reaching adults with growing need. These groups were small (38 discontinued, 25 initiated), so these comparisons are imprecise.

This study has several strengths. First, we used individual-level longitudinal data spanning early implementation, augmenting the existing literature that has primarily relied on administrative caseload data, aggregate food-pantry utilization, or repeated cross-sectional surveys.^20–24,40,41^ The cohort design allows us to observe within-person change over time rather than inferring ecologically from population aggregates. Second, by combining 12-month and 30-day SNAP measures, we could distinguish adults who continued, discontinued or initiated SNAP between assessments.

The following limitations also apply. First, this was not a causal test of OBBBA on SNAP receipt and food insecurity, and it is too early to conduct one. The time period we examined spans several concurrent events (end of ABAWD waivers, shutdown-related benefit interruption, pre-existing national SNAP decline) that could affect the outcomes under study. Second, our binary classification of states as early-versus not-yet-implementing was based on a single point in time and collapses important differences among not-yet-implementing states (which include both high-cost, high-safety-net states such as California and lower-cost, more rural states such as Montana and North Dakota). Third, we did not apply a correction for multiple comparisons, and findings should be treated as descriptive and hypothesis-generating. Fourth, we approximated SNAP eligibility using household income below 200% FPG at a single point in time; income categories are coarse and can change over the year. Fifth, SNAP receipt was self-reported, and the groups who discontinued or initiated SNAP were small, limiting precision. Because the 12-month food insecurity window at March 2026 overlaps the period in which SNAP status changed, we could not determine whether food insecurity preceded or followed discontinuing or initiating SNAP.

## CONCLUSION

In the first months of OBBBA implementation, food insecurity remained high and did not improve among low-income adults. Changes did not yet differ between early- and not-yet-implementing states. Policies that keep benefits flowing for eligible people, including those with continued food insecurity, such as extended certification periods, simplified recertification, and protection against benefit interruptions, may limit harm as implementation proceeds.^31^

## Conflict of interest

All authors declare no conflicts of interest. The City University of New York (CUNY) School of Public Health received research support from Pfizer during the conduct of this study, which was paid directly to CUNY, with Dr. Nash serving as the Principal Investigator.

## Acknowledgements

We thank the participants of the Communities, Households, and SARS-CoV-2 Epidemiology COVID Cohort Study for their contributions to the advancement of science.

During the preparation of this work, the authors used the AI tool Undermind as part of the process to identify relevant literature. All papers identified with Undermind and cited in the text were reviewed by the authors.

The authors also used Claude.ai (Anthropic) in an iterative and author-directed capacity to develop and refine R and SAS code for statistical analyses and to aid with creating reproducible and fully formatted tables. Code was first generated by the authors without Claude assistance to create variables and confirm sample sizes and frequencies. All analytic decisions including outcome definitions, covariate selections, model specification and interpretation of results were made by authors. The authors reviewed and edited all AI-assisted code for accuracy, and the authors take full responsibility for the content of the published article.

## Data availability statement

The datasets generated and/or analyzed during the current study are not publicly available, but are available from the authors upon reasonable request, subject to available data management resources.

## Funding

This work was supported by The Schmidt Family Foundation, the CUNY Institute for Implementation Science in Population Health (cunyisph.org). The CHASING COVID Cohort has also been supported by the National Institute of Allergy and Infectious Diseases (NIAID) award number UH3AI133675 (MPIs: D Nash and C Grov), NIMH award RF1MH132360 (MPIs: D Nash and A Parcesepe), National Institute of Child Health and Human Development grant P2C HD050924 (Carolina Population Center), Pfizer Inc., and the COVID-19 Grant Program of the CUNY Graduate School of Public Health and Health Policy. The funders played no role in the production of this manuscript or endorsed findings.

---

**Supplemental Figure 1.**
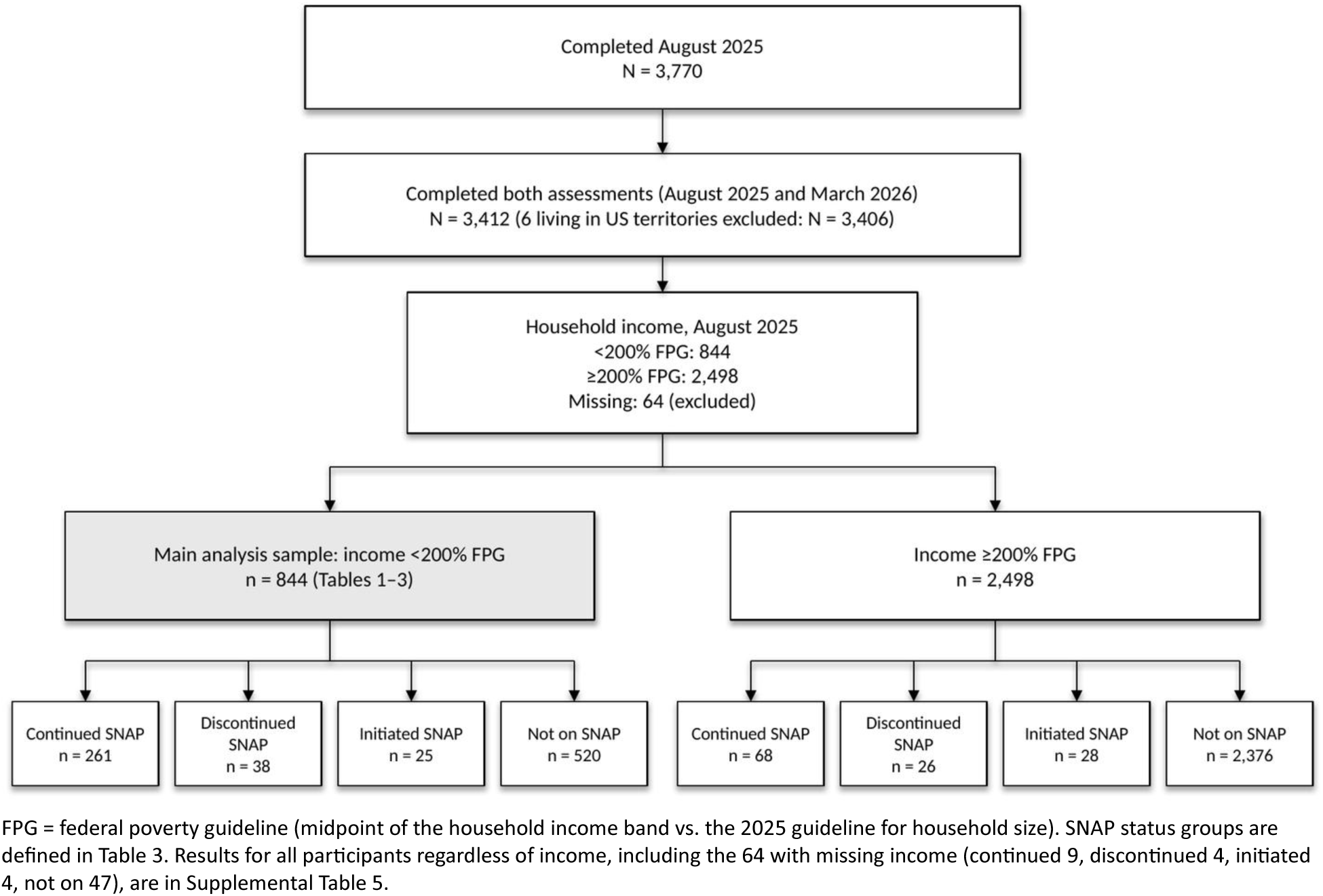
Participant Flow. FPG = federal poverty guideline (midpoint of the household income band vs. the 2025 guideline for household size). SNAP status groups are defined in Table 3. Results for all participants regardless of income, including the 64 with missing income (continued 9, discontinued 4, initiated 4, not on 47), are in Supplemental Table 5.

**Supplemental Table 1.** Early-Implementing versus Not-Yet-Implementing States.

| State Group | State (March 2026) |
|---|---|
| Early-implementing | AK |
| Early-implementing | AL |
| Early-implementing | AR |
| Early-implementing | AZ |
| Early-implementing | CO |
| Early-implementing | CT |
| Early-implementing | DE |
| Early-implementing | FL |
| Early-implementing | GA |
| Early-implementing | HI |
| Early-implementing | IA |
| Early-implementing | ID |
| Early-implementing | IN |
| Early-implementing | KS |
| Early-implementing | KY |
| Early-implementing | LA |
| Early-implementing | MA |
| Early-implementing | MD |
| Early-implementing | ME |
| Early-implementing | MO |
| Early-implementing | MS |
| Early-implementing | NC |
| Early-implementing | NE |
| Early-implementing | NH |
| Early-implementing | NM |
| Early-implementing | OH |
| Early-implementing | OK |
| Early-implementing | PA |
| Early-implementing | RI |
| Early-implementing | SC |
| Early-implementing | SD |
| Early-implementing | TN |
| Early-implementing | TX |
| Early-implementing | UT |
| Early-implementing | VA |
| Early-implementing | VT |
| Early-implementing | WI |
| Early-implementing | WV |
| Early-implementing | WY |
| Not-yet-implementing | CA |
| Not-yet-implementing | DC |
| Not-yet-implementing | IL |
| Not-yet-implementing | MI |
| Not-yet-implementing | MN |
| Not-yet-implementing | MT |
| Not-yet-implementing | ND |
| Not-yet-implementing | NJ |
| Not-yet-implementing | NV |
| Not-yet-implementing | NY |
| Not-yet-implementing | OR |
| Not-yet-implementing | WA |

**Supplemental Table 2.** Discontinuing and Initiating SNAP Between August 2025 and March 2026 by State ABAWD Implementation Timing, Among Adults With Household Income <200% FPG.

| **Measure** | **Overall, n/N (%)** | **Early-implementing, n/N (%)** | **Not-yet-implementing, n/N (%)** | **RR (95% CI)** | **p** |
|---|---|---|---|---|---|
| Discontinued SNAP, among those with SNAP in the past 12 months at March 2026 | 38/324 (11.7%) | 24/181 (13.3%) | 13/140 (9.3%) | 1.43 (0.75–2.70) | 0.27 |
| Initiated SNAP, among those with no SNAP in the past 12 months at August 2025 | 25/503 (5.0%) | 16/335 (4.8%) | 9/159 (5.7%) | 0.84 (0.38–1.87) | 0.68 |

Adults with household income <200% FPG in August 2025 (N = 844). Overall includes 12 participants with missing state. RR from modified Poisson regression with robust standard errors; reference = not-yet-implementing states.

**Supplemental Table 3.** Characteristics of Adults With Household Income <200% FPG, by SNAP Status Between August 2025 and March 2026.

| Characteristic | Continued SNAP, N = 261 | Discontinued SNAP, N = 38 | Initiated SNAP, N = 25 | Not on SNAP, N = 520 | Overall, N = 844 |
|---|---|---|---|---|---|
| **Household income, % of federal poverty guideline (August 2025)** | 83 (46, 113) | 96 (54, 160) | 113 (70, 142) | 132 (83, 166) | 112 (70, 160) |
| **Age group** |  |  |  |  |  |
| 18-34 | 50 (19.2%) | 11 (28.9%) | 8 (32.0%) | 131 (25.2%) | 200 (23.7%) |
| 35-44 | 81 (31.0%) | 17 (44.7%) | 11 (44.0%) | 134 (25.8%) | 243 (28.8%) |
| 45-54 | 49 (18.8%) | 8 (21.1%) | 2 (8.0%) | 98 (18.8%) | 157 (18.6%) |
| 55-64 | 36 (13.8%) | 2 (5.3%) | 1 (4.0%) | 65 (12.5%) | 104 (12.3%) |
| 65+ | 45 (17.2%) | 0 (0.0%) | 3 (12.0%) | 92 (17.7%) | 140 (16.6%) |
| **Gender** |  |  |  |  |  |
| Female | 180 (69.0%) | 26 (68.4%) | 14 (56.0%) | 336 (64.6%) | 556 (65.9%) |
| Male | 73 (28.0%) | 9 (23.7%) | 10 (40.0%) | 162 (31.2%) | 254 (30.1%) |
| Non-binary | 8 (3.1%) | 3 (7.9%) | 1 (4.0%) | 22 (4.2%) | 34 (4.0%) |
| **Race/ethnicity** |  |  |  |  |  |
| Asian/Pacific Islander | 4 (1.5%) | 2 (5.3%) | 1 (4.0%) | 29 (5.6%) | 36 (4.3%) |
| Black non-Hispanic | 57 (21.8%) | 5 (13.2%) | 6 (24.0%) | 82 (15.8%) | 150 (17.8%) |
| Hispanic | 65 (24.9%) | 15 (39.5%) | 8 (32.0%) | 123 (23.7%) | 211 (25.0%) |
| White non-Hispanic | 120 (46.0%) | 14 (36.8%) | 10 (40.0%) | 257 (49.4%) | 401 (47.5%) |
| Other/Multiracial | 15 (5.7%) | 2 (5.3%) | 0 (0.0%) | 28 (5.4%) | 45 (5.3%) |
| Unknown | 0 (0.0%) | 0 (0.0%) | 0 (0.0%) | 1 (0.2%) | 1 (0.1%) |
| **Sexual orientation** |  |  |  |  |  |
| Bisexual | 19 (7.3%) | 3 (7.9%) | 3 (12.0%) | 47 (9.0%) | 72 (8.5%) |
| Lesbian or Gay | 38 (14.6%) | 3 (7.9%) | 1 (4.0%) | 65 (12.5%) | 107 (12.7%) |
| Straight | 162 (62.1%) | 24 (63.2%) | 16 (64.0%) | 317 (61.0%) | 519 (61.5%) |
| Other | 6 (2.3%) | 1 (2.6%) | 0 (0.0%) | 11 (2.1%) | 18 (2.1%) |
| Unknown | 36 (13.8%) | 7 (18.4%) | 5 (20.0%) | 80 (15.4%) | 128 (15.2%) |
| **Education** |  |  |  |  |  |
| ≤High school | 88 (33.7%) | 16 (42.1%) | 12 (48.0%) | 118 (22.7%) | 234 (27.7%) |
| Some college | 113 (43.3%) | 9 (23.7%) | 9 (36.0%) | 206 (39.6%) | 337 (39.9%) |
| College graduate | 60 (23.0%) | 13 (34.2%) | 4 (16.0%) | 196 (37.7%) | 273 (32.3%) |
| **Employment status** |  |  |  |  |  |
| Employed | 90 (34.5%) | 25 (65.8%) | 17 (68.0%) | 279 (53.7%) | 411 (48.7%) |
| Out of work | 46 (17.6%) | 4 (10.5%) | 1 (4.0%) | 51 (9.8%) | 102 (12.1%) |
| Homemaker | 17 (6.5%) | 3 (7.9%) | 2 (8.0%) | 40 (7.7%) | 62 (7.3%) |
| Student | 13 (5.0%) | 3 (7.9%) | 2 (8.0%) | 30 (5.8%) | 48 (5.7%) |
| Retired | 45 (17.2%) | 0 (0.0%) | 2 (8.0%) | 90 (17.3%) | 137 (16.2%) |
| Unable to work | 50 (19.2%) | 3 (7.9%) | 1 (4.0%) | 30 (5.8%) | 84 (10.0%) |
| **Household income category** |  |  |  |  |  |
| <\$25,000 | 192 (73.6%) | 17 (44.7%) | 6 (24.0%) | 194 (37.3%) | 409 (48.5%) |
| \$25,000-<\$50,000 | 54 (20.7%) | 13 (34.2%) | 15 (60.0%) | 230 (44.2%) | 312 (37.0%) |
| \$50,000-<\$100,000 | 15 (5.7%) | 8 (21.1%) | 4 (16.0%) | 93 (17.9%) | 120 (14.2%) |
| \$100,000 or more | 0 (0.0%) | 0 (0.0%) | 0 (0.0%) | 3 (0.6%) | 3 (0.4%) |
| **Marital status** |  |  |  |  |  |
| Married/partnered | 52 (19.9%) | 19 (50.0%) | 8 (32.0%) | 195 (37.5%) | 274 (32.5%) |
| Previously married | 81 (31.0%) | 6 (15.8%) | 8 (32.0%) | 122 (23.5%) | 217 (25.7%) |
| Never married | 127 (48.7%) | 13 (34.2%) | 9 (36.0%) | 203 (39.0%) | 352 (41.7%) |
| **Any child <18 in household** |  |  |  |  |  |
| Yes | 92 (35.2%) | 16 (42.1%) | 11 (44.0%) | 161 (31.0%) | 280 (33.2%) |
| No | 169 (64.8%) | 22 (57.9%) | 14 (56.0%) | 358 (68.8%) | 563 (66.7%) |
| **Single parent** |  |  |  |  |  |
| Yes | 57 (21.8%) | 3 (7.9%) | 6 (24.0%) | 57 (11.0%) | 123 (14.6%) |
| No | 35 (13.4%) | 13 (34.2%) | 5 (20.0%) | 104 (20.0%) | 157 (18.6%) |
| No children in household | 169 (64.8%) | 22 (57.9%) | 14 (56.0%) | 358 (68.8%) | 563 (66.7%) |
| **Parent of child <14 (SNAP work-requirement exemption threshold)** |  |  |  |  |  |
| Yes | 64 (24.5%) | 13 (34.2%) | 6 (24.0%) | 106 (20.4%) | 189 (22.4%) |
| No | 28 (10.7%) | 3 (7.9%) | 5 (20.0%) | 55 (10.6%) | 91 (10.8%) |
| **Any disability (ACS 6-item screener)** |  |  |  |  |  |
| Yes | 128 (49.0%) | 15 (39.5%) | 4 (16.0%) | 161 (31.0%) | 308 (36.5%) |
| No | 133 (51.0%) | 23 (60.5%) | 21 (84.0%) | 359 (69.0%) | 536 (63.5%) |
| **SNAP work requirement (likely subject vs. exempt)** |  |  |  |  |  |
| Likely subject | 70 (26.8%) | 16 (42.1%) | 14 (56.0%) | 224 (43.1%) | 324 (38.4%) |
| Exempt | 191 (73.2%) | 22 (57.9%) | 11 (44.0%) | 295 (56.7%) | 519 (61.5%) |
| Missing | 0 (0.0%) | 0 (0.0%) | 0 (0.0%) | 1 (0.2%) | 1 (0.1%) |
| **State SNAP ABAWD implementation timing** |  |  |  |  |  |
| Early-implementing | 141 (54.0%) | 24 (63.2%) | 16 (64.0%) | 343 (66.0%) | 524 (62.1%) |
| Not-yet-implementing | 118 (45.2%) | 13 (34.2%) | 9 (36.0%) | 168 (32.3%) | 308 (36.5%) |
| Missing state | 2 (0.8%) | 1 (2.6%) | 0 (0.0%) | 9 (1.7%) | 12 (1.4%) |
| **Employed, Mar 2026** | 87 (33.3%) | 22 (57.9%) | 16 (64.0%) | 298 (57.3%) | 423 (50.1%) |
| **Household income, Mar 2026** |  |  |  |  |  |
| <200% FPG | 244 (93.5%) | 32 (84.2%) | 24 (96.0%) | 414 (79.6%) | 714 (84.6%) |
| ≥200% FPG | 13 (5.0%) | 5 (13.2%) | 0 (0.0%) | 92 (17.7%) | 110 (13.0%) |
| Missing | 4 (1.5%) | 1 (2.6%) | 1 (4.0%) | 14 (2.7%) | 20 (2.4%) |

Adults with household income <200% FPG in August 2025. SNAP status groups are defined in Table 3. Characteristics measured in August 2025 unless noted. Median (Q1, Q3) or n (%), column percentages.

**Supplemental Table 4.** Change in Past 12-Month Outcomes from August 2025 to March 2026 Overall and by State ABAWD Implementation Timing, Among All Participants Regardless of Income (N = 3,406)

| Outcome | State group | N | Aug 2025, n (%) | Mar 2026, n (%) | RR (95% CI) | Ratio of RRs, early- vs. not-yet-implementing (95% CI) | p |
|---|---|---|---|---|---|---|---|
| **All income levels** |  |  |  |  |  |  |  |
| SNAP receipt | Early-implementing | 1,952 | 270 (13.8%) | 247 (12.7%) | 0.91 (0.85–0.99) | 0.95 (0.86–1.06) | 0.39 |
|  | Not-yet-implementing | 1,412 | 221 (15.7%) | 212 (15.0%) | 0.96 (0.89–1.04) |  |  |
|  | **Overall** | 3,406 | 495 (14.5%) | 463 (13.6%) | **0.94 (0.89–0.99)** |  |  |
| Food insecurity | Early-implementing | 1,952 | 463 (23.7%) | 486 (24.9%) | 1.05 (0.99–1.11) | 1.00 (0.91–1.11) | 0.94 |
|  | Not-yet-implementing | 1,412 | 284 (20.1%) | 297 (21.0%) | 1.05 (0.96–1.14) |  |  |
|  | **Overall** | 3,406 | 756 (22.2%) | 792 (23.3%) | **1.05 (1.00–1.10)** |  |  |
| Employed | Early-implementing | 1,952 | 1,334 (68.3%) | 1,327 (68.0%) | 0.99 (0.98–1.01) | 0.99 (0.97–1.01) | 0.42 |
|  | Not-yet-implementing | 1,412 | 999 (70.8%) | 1,004 (71.1%) | 1.01 (0.99–1.02) |  |  |
|  | **Overall** | 3,406 | 2,365 (69.4%) | 2,363 (69.4%) | 1.00 (0.99–1.01) |  |  |
| Any utility/bill hardship | Early-implementing | 1,952 | 339 (17.4%) | 316 (16.2%) | **0.93 (0.86–1.02)** | 0.95 (0.82–1.10) | 0.51 |
|  | Not-yet-implementing | 1,412 | 192 (13.6%) | 188 (13.3%) | 0.98 (0.87–1.10) |  |  |
|  | **Overall** | 3,406 | 535 (15.7%) | 507 (14.9%) | **0.95 (0.88–1.02)** |  |  |
| Housing instability | Early-implementing | 1,952 | 747 (38.3%) | 739 (37.9%) | 0.99 (0.93–1.05) | 0.96 (0.87–1.05) | 0.34 |
|  | Not-yet-implementing | 1,412 | 510 (36.1%) | 528 (37.4%) | 1.04 (0.97–1.11) |  |  |
|  | **Overall** | 3,406 | 1,275 (37.4%) | 1,283 (37.7%) | 1.01 (0.96–1.05) |  |  |

All participants who completed both assessments, regardless of income (N = 3,406; includes 64 with missing income and 42 with missing state, who are in Overall only). Modified Poisson generalized estimating equations (log link, exchangeable correlation, clustered on participant). RR = March 2026 vs. August 2025 within each state group. Ratio of RRs = wave × state-group interaction (difference-in-differences on the ratio scale), reference = not-yet-implementing states; p = p for interaction. IPCW-weighted estimates were nearly identical (not shown). Item nonresponse was ≤0.8% for each outcome at either assessment (coded No). SNAP receipt = past 12 months; food insecurity = Hunger Vital Sign.

**Supplemental Table 5.**
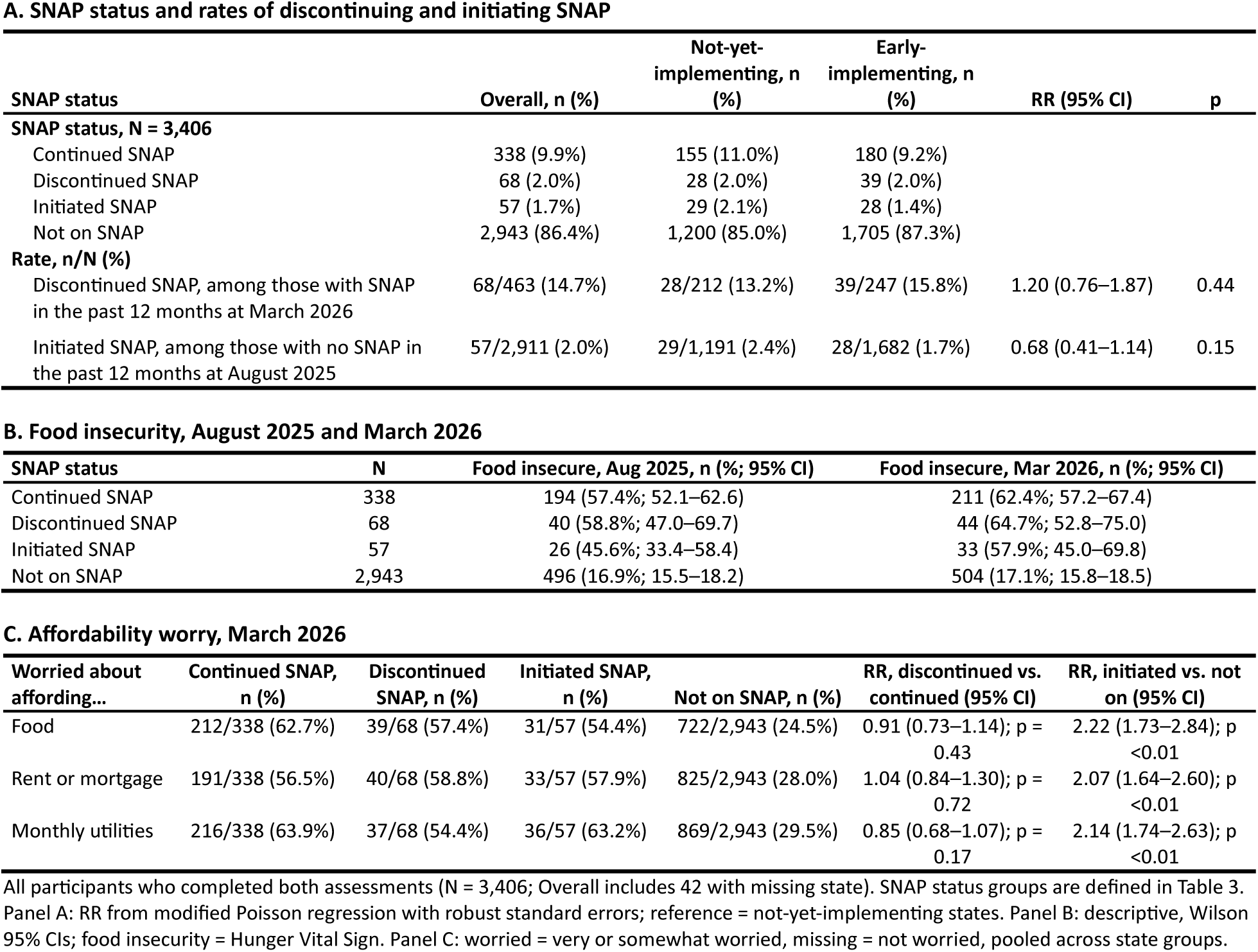
SNAP Status Between August 2025 and March 2026, Food Insecurity, and Affordability Worry Among All Participants Regardless of Income, by State ABAWD Implementation Timing.

| **A. SNAP status and rates of discontinuing and initiating SNAP** |  |  |  |  |  |  |
| **SNAP status** | **Overall, n (%)** | **Not-yet-implementing, n (%)** | **Early-implementing, n (%)** | **RR (95% CI)** | **p** |  |
|---|---|---|---|---|---|---|
| **SNAP status, N = 3,406** |  |  |  |  |  |  |
| Continued SNAP | 338 (9.9%) | 155 (11.0%) | 180 (9.2%) |  |  |  |
| Discontinued SNAP | 68 (2.0%) | 28 (2.0%) | 39 (2.0%) |  |  |  |
| Initiated SNAP | 57 (1.7%) | 29 (2.1%) | 28 (1.4%) |  |  |  |
| Not on SNAP | 2,943 (86.4%) | 1,200 (85.0%) | 1,705 (87.3%) |  |  |  |
| **Rate, n/N (%)** |  |  |  |  |  |  |
| Discontinued SNAP, among those with SNAP in the past 12 months at March 2026 | 68/463 (14.7%) | 28/212 (13.2%) | 39/247 (15.8%) | 1.20 (0.76–1.87) | 0.44 |  |
| Initiated SNAP, among those with no SNAP in the past 12 months at August 2025 | 57/2,911 (2.0%) | 29/1,191 (2.4%) | 28/1,682 (1.7%) | 0.68 (0.41–1.14) | 0.15 |  |
| **B. Food insecurity, August 2025 and March 2026** |  |  |  |  |  |  |
| **SNAP status** | **N** | **Food insecure, Aug 2025, n (%; 95% CI)** | **Food insecure, Mar 2026, n (%; 95% CI)** |  |  |  |
| Continued SNAP | 338 | 194 (57.4%; 52.1–62.6) | 211 (62.4%; 57.2–67.4) |  |  |  |
| Discontinued SNAP | 68 | 40 (58.8%; 47.0–69.7) | 44 (64.7%; 52.8–75.0) |  |  |  |
| Initiated SNAP | 57 | 26 (45.6%; 33.4–58.4) | 33 (57.9%; 45.0–69.8) |  |  |  |
| Not on SNAP | 2,943 | 496 (16.9%; 15.5–18.2) | 504 (17.1%; 15.8–18.5) |  |  |  |
| **C. Affordability worry, March 2026** |  |  |  |  |  |  |
| **Worried about affording…** | **Continued SNAP, n (%)** | **Discontinued SNAP, n (%)** | **Initiated SNAP, n (%)** | **Not on SNAP, n (%)** | **RR, discontinued vs. continued (95% CI)** | **RR, initiated vs. not on (95% CI)** |
| Food | 212/338 (62.7%) | 39/68 (57.4%) | 31/57 (54.4%) | 722/2,943 (24.5%) | 0.91 (0.73–1.14); p = 0.43 | 2.22 (1.73–2.84); p <0.01 |
| Rent or mortgage | 191/338 (56.5%) | 40/68 (58.8%) | 33/57 (57.9%) | 825/2,943 (28.0%) | 1.04 (0.84–1.30); p = 0.72 | 2.07 (1.64–2.60); p <0.01 |
| Monthly utilities | 216/338 (63.9%) | 37/68 (54.4%) | 36/57 (63.2%) | 869/2,943 (29.5%) | 0.85 (0.68–1.07); p = 0.17 | 2.14 (1.74–2.63); p <0.01 |

All participants who completed both assessments (N = 3,406; Overall includes 42 with missing state). SNAP status groups are defined in Table 3. Panel A: RR from modified Poisson regression with robust standard errors; reference = not-yet-implementing states. Panel B: descriptive, Wilson 95% CIs; food insecurity = Hunger Vital Sign. Panel C: worried = very or somewhat worried, missing = not worried, pooled across state groups.

**Supplemental Table 6.** SNAP Receipt and Food Insecurity Among Adults With Household Income <200% FPG Likely Subject to vs. Exempt From the SNAP Work Requirement, by State ABAWD Implementation Timing.

| Outcome / state group | Work requirement | N | Aug 2025, n (%) | Mar 2026, n (%) | RR (95% CI) | Subject vs. exempt, ratio of RRs (95% CI) | Triple difference (95% CI) |
|---|---|---|---|---|---|---|---|
| **SNAP receipt** |  |  |  |  |  |  |  |
| Early-implementing | Subject | 197 | 54 (27.4%) | 52 (26.4%) | 0.96 (0.78–1.19) | 1.01 (0.81–1.26) | 0.89 (0.68–1.16) |
|  | Exempt | 327 | 135 (41.3%) | 129 (39.4%) | 0.96 (0.89–1.03) |  |  |
| Not-yet-implementing | Subject | 124 | 46 (37.1%) | 47 (37.9%) | 1.02 (0.91–1.14) | 1.13 (0.98–1.31) |  |
|  | Exempt | 183 | 103 (56.3%) | 93 (50.8%) | **0.90 (0.82–0.99)** |  |  |
| **Food insecurity (Hunger Vital Sign)** |  |  |  |  |  |  |  |
| Early-implementing | Subject | 197 | 99 (50.3%) | 110 (55.8%) | 1.11 (0.99–1.25) | 1.10 (0.96–1.26) | 0.96 (0.72–1.26) |
|  | Exempt | 327 | 166 (50.8%) | 168 (51.4%) | 1.01 (0.94–1.09) |  |  |
| Not-yet-implementing | Subject | 124 | 41 (33.1%) | 49 (39.5%) | 1.20 (0.96–1.48) | 1.15 (0.90–1.46) |  |
|  | Exempt | 183 | 96 (52.5%) | 100 (54.6%) | 1.04 (0.93–1.17) |  |  |

Subject = aged 18–64, no child <14, no disability, not pregnant (August 2025); exempt = all others. Hours worked were not measured, so some subject adults may already meet the work requirement. Modified Poisson generalized estimating equations (exchangeable correlation, clustered on participant). Ratio of RRs = change among subject adults divided by change among exempt adults in the same state group. Triple difference = that ratio in early-implementing states divided by the ratio in not-yet-implementing states (1.00 = no difference). IPCW-weighted estimates were nearly identical (not shown).

## APPENDIX 1. IPCW

To correct for loss to follow-up at the March 2026 assessment, we constructed stabilized inverse probability of censoring weights (IPCW) truncated at the 1st and 99th percentiles to limit the influence of extreme weights on variance. The final logistic regression model included sociodemographic variables (age, gender, race/ethnicity, sexual orientation, education, employment, income, marital status, single-parent status, disability, food insecurity, housing instability) plus GAD-7 and PHQ-8 continuous symptom scores. A small number of respondents (n=7) completed both assessments per the completion flag but provided almost no substantive item-level responses and were excluded from model fitting and instead assigned a neutral weight (=1), since a completion probability could not be meaningfully estimated given near-absent covariate data. Standard maximum-likelihood logistic regression produced quasi-complete separation, evidenced by implausibly large coefficients (magnitude ∼11) with correspondingly enormous standard errors (∼265) on several sparse categories; we resolved this by refitting with Firth’s penalized (bias-reduced) logistic regression.

## Notes

### Competing Interest Statement

The authors have declared no competing interest.

